# Novel Insights into Salt-Sensitivity of Blood Pressure in African Adults with and without HIV: Comprehensive Inflammatory, renal and Cardiometabolic Profiling in a Zambian Cohort

**DOI:** 10.1101/2025.11.17.25340449

**Authors:** Sepiso K. Masenga, Joreen P. Povia, Catherine Anna-Marie Graham, Yiannis Mavrommatis, Benson M. Hamooya, Leta Pilic, Annet Kirabo

## Abstract

**Background:** Salt sensitivity of blood pressure (SSBP) amplifies cardiovascular risk in hypertension. Although we previously found that SSBP was more prevalent in people with HIV (PWH) compared to the HIV-uninfected, yet its determinants in PWH remain understudied in sub-Saharan Africa. In this study, we hypothesized that chronic inflammation and cardiometabolic disturbances would be uniquely associated with SSBP in Zambian adults with and without HIV.

**Methods:** We performed a cross-sectional SSBP assessment in 366 adults (269 PWH, 97 without HIV [PWTH]). We also performed echocardiography, carotid ultrasonography, flow-mediated dilation, lipid profiles, renal function, inflammatory markers, taste perception, and 24-hour dietary recalls. SSBP was defined as a ≥10mmHg mean arterial pressure increase between high-salt and low-salt intervention. Multivariate logistic regression models were used to identify independent correlates of SSBP in the overall population (adjusted for age, sex, and HIV status) and separately in PWH and PWTH (adjusted for age and sex).

**Results:** In the overall population multivariate analysis, hypertension (AOR=12.28), IPROS (AOR=5.70), peripheral artery disease (AOR=2.63), left ventricular hypertrophy (AOR=2.31), and higher cardiovascular risk scores were independently associated with SSBP. PWH were older (49 ± 12 vs. 44 ± 17 years, p= 0.015) and exhibited lower BMI (25.04 vs. 26.44 kg/m², p=0.045) and waist circumference (83.79 vs. 87.76 cm, p= 0.037) than PWTH. PWH demonstrated elevated salt-taste recognition thresholds (0.467 vs. 0.233 g/0.5L, p= 0.0014) and dysregulated metabolic/inflammatory profiles, including higher triglycerides, d-dimer, high-sensitivity C-reactive protein (hs-CRP), IL-6, and hormonal profile including renin, aldosterone, angiotensin II, and N-terminal pro-B-type natriuretic peptide (NT-proBNP) (p< 0.05). Hypertension strongly predicted SSBP in both groups in univariate (PWH: OR = 13.08, 95% CI 6.62–25.81; PWTH: OR = 28.33, 95% CI 7.69–104.35, p<0.001) and multivariate analysis (PWH: AOR = 10.27, 95% CI 5.02–21.00; PWTH: AOR = 24.68, 95% CI 5.45–111.73, P<0.0001). In multivariate analysis, an immediate pressor response to oral salt (IPROS) was independently associated with SSBP in PWH (AOR = 19.90, 95% CI 6.61–59.91) and PWTH (AOR = 18.49, 95% CI 2.14–159.28). In a comprehensive multivariate model, larger waist circumference was associated with reduced odds of SSBP (AOR=0.94 per cm, p=0.016), while greater left ventricular posterior wall thickness was associated with increased odds (AOR=21.44, p=0.027) among PWH. Left ventricular mass index, atherosclerotic cardiovascular disease (ASCVD) risk and plasma creatinine were additional correlates in PWH (p< 0.01) but not in PWTH. In PWTH, atrial natriuretic peptide (AOR=1.00, 95% CI 1.00-1.00, p=0.021) and peripheral artery disease (AOR= 5.39, 95% CI 1.20-24.06, p=0.027) were the only unique factors associated with SSBP compared to PWH.

**Conclusion:** SSBP in this Zambian cohort is associated with a complex interplay of traditional and HIV-specific factors. The strong independent association of IPROS with SSBP across all analyses supports its potential utility as a clinical screening tool. The distinct correlates in PWH, particularly the prominent role of cardiac structural changes and the unexpected association with marital status, highlight the need for HIV-specific approaches to salt sensitivity assessment and management.

## 1 Introduction

Salt sensitivity of blood pressure (SSBP), an exaggerated blood pressure response to salt intake, is an established risk factor for hypertension, cardiovascular end-organ damage, and mortality ^1^. However, most SSBP studies have been conducted in Western countries and predominantly in White or non-HIV populations. We previously reported that SSBP prevalence in Black individuals in sub-Saharan Africa is as high as 75–80%, similar to western cohorts ^2^. Intriguingly, SSBP disproportionately affects groups prone to hypertension such as the elderly, Black individuals, and women ^1^, but its mechanisms extend beyond traditional renal sodium handling to involve the immune system and inflammation ^3–5^. High salt intake itself can initiate inflammatory processes that raise blood pressure, and conversely chronic inflammation may impair renal sodium excretion, creating a vicious cycle of salt-sensitive hypertension ^3–5^.

People living with HIV (PWH) represent a population at heightened risk for cardiovascular disease (CVD). HIV infection and long-term antiretroviral therapy (ART) are associated with a 1.5–2-fold increased risk of hypertension and CVD compared to HIV-negative individuals ^6,7^. Even with viral suppression on ART, persistent immune activation and inflammation remain hallmarks of chronic HIV infection ^8^. Elevated levels of interleukin-6 (IL-6), C-reactive protein (CRP), and other cytokines are common in treated HIV and strongly predict CVD events and mortality ^8^. It has been postulated that this chronic inflammatory milieu, alongside traditional risk factors, contributes to greater hypertension in PWH ^9–12^. Recent evidence suggests salt sensitivity might be especially prevalent in PLWH. For example, we have previously observed that a disproportionate number of HIV-positive hypertensive Zambian adults had SSBP, significantly higher than in HIV-negative hypertensives ^13^. Moreover, HIV-positive hypertensives in that study were more likely to exhibit a non-dipping blood pressure pattern at night ^13^, compounding their cardiovascular risk.

Despite these hints, no prior study in Africa has comprehensively phenotyped salt sensitivity in an HIV-positive cohort with concurrent assessment of inflammatory and metabolic profiles. We hypothesized that in PWH, chronic inflammation and metabolic disturbances would be uniquely associated with SSBP, helping to explain the excess hypertension risk in this population. To test this, we conducted a cross-sectional study in Zambian adults with HIV, performing detailed cardiovascular, metabolic, and inflammatory profiling. Here we present novel findings on the relationships between SSBP, HIV-related inflammation, and cardiometabolic factors, positioning this work as the first in an African HIV cohort to investigate these associations. Our results provide insight into mechanisms of HIV-associated hypertension and suggest potential targets for CVD prevention in this high-risk group.

## 2 Methods

### 2.1 Study design and participants

This cross-sectional study recruited 366 adults (269 PWH, 97 PWTH) from a cohort of individuals with known SSBP status ^14^ who attended routine medical checkup at a tertiary hospital in Livingstone, Zambia, **Figure 1A**.

**Figure 1.**
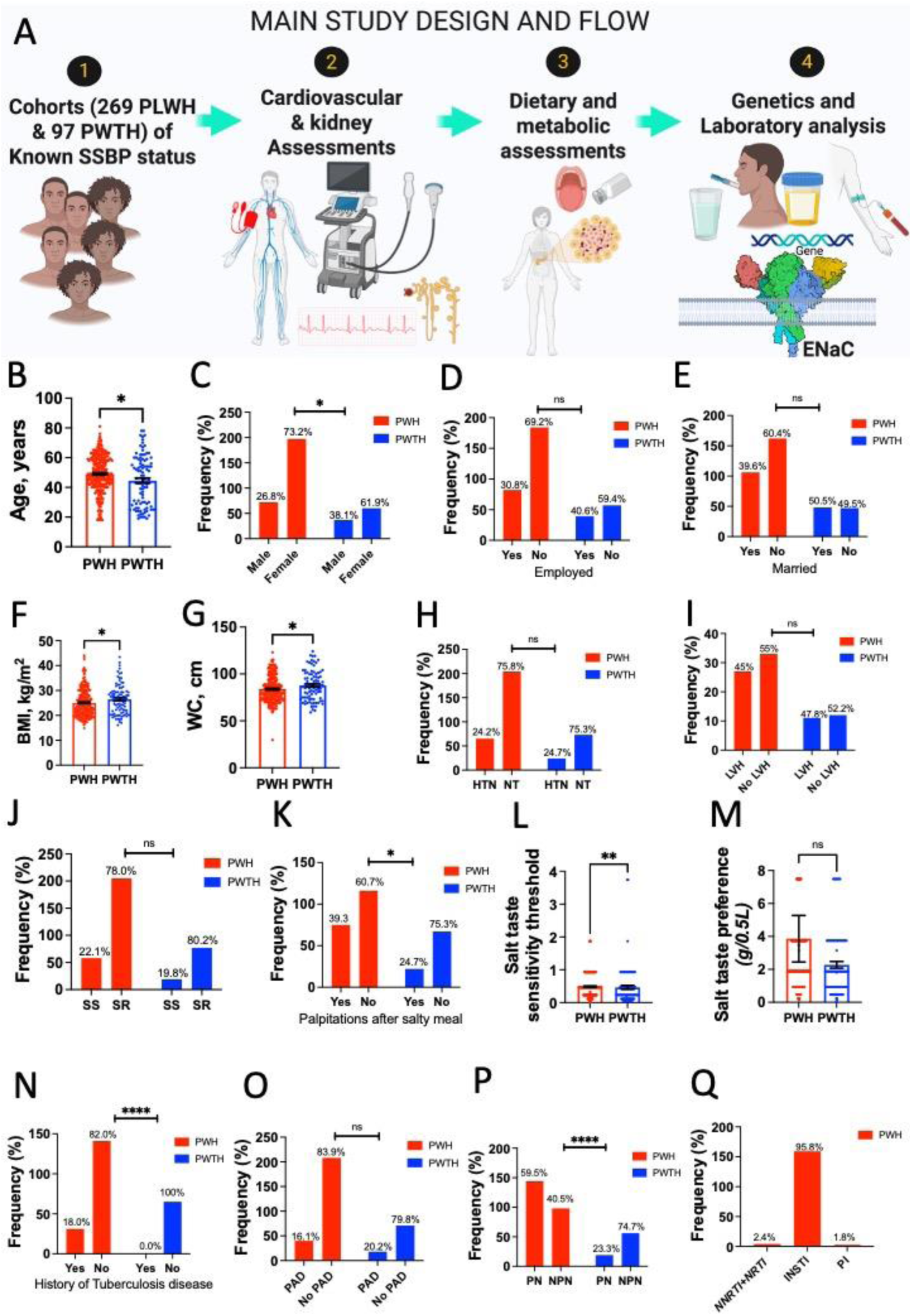
General characteristics of the study population. (A) A total of 366 participants including 269 persons with HIV (PLWH or PWH) and 97 persons without HIV (PWTH) with known salt sensitivity of blood pressure (SSBP) status cardiovascular, dietary and laboratory characteristics were recruited. (B) PWH were slightly older than PWTH with mean age of 49±12 vs. 44±17 years, respectively, p=0.015, welch’s t test. (C) Among PWH 26.8% were males and 73.2% were females compared to 38.1 and 61.9% among PWTH. Proportions of those (D) employed and (E) married are presented. PWH had lower (F) body mass index (BMI) compared to PWTH (25.04 vs. 26.44 kg/m^2^, p=0.045, welch’s t test) and this was similar for (G) waist circumference (83.79 vs 87.76 cm, p=0.037). Proportions of participants with (H) hypertension (HTN) or normotensive (NT), (I) left ventricular hypertrophy (LVH) and (J) salt-sensitive or salt resistant were not significantly different between PWH and PWTH except for participants who had a (K) history of feeling heart palpitations after consuming a salty meal. PWH had a higher (L) salt-taste sensitivity threshold compared to PWHT (0.467 vs. 0.233 g/0.5L, p=0.0014, Mann-Whitney test), and consequently PWH also had a higher (M) salt taste preference compared to PWTH (3.85 vs 2.27 g/0.5L, p=0.267) although not statistically significant. (N) history of tuberculosis and (P) peripheral neuropathy was associated with HIV status but this was not the case with (O) peripheral artery disease. The majority of PWH were on (Q) integrase strand inhibitor-based antiretroviral therapy.

### 2.2 Eligibility

Adults aged ≥18 years with confirmed HIV infection on stable ART for ≥6 months and virologically suppressed (viral load <50 copies/mL) were recruited during routine clinic visits. PWTH had negative HIV tests. We included both normotensive and hypertensive PWH and PWTH, excluding those with diabetes, pregnancy, severe renal/hepatic disease, active opportunistic infections, or other serious comorbidities to avoid confounding influences on inflammation.

### 2.3 Demographic data

We used Research electronic data capture (REDCap) to create standardized questionnaires that we used to capture demographic data such as age, sex, employment, marital status, and medical history.

### 2.4 Anthropometric measurements

Height (m) and weight (kg) were measured using a stadiometer and body mass index (BMI) were calculated using the equation kg/m^2^. Waist circumference was measured midway between the iliac crest and lower rib.

### 2.5 Dietary assessments

#### 2.5.1 Taste threshold

To assess salt taste threshold, seven sodium chloride solutions (0.0, 5.0, 15.0, 30.0, 60.0, 120.0 and 240.0 g/L) were freshly prepared in spring water on the day of testing. These were administered to participants in ascending order of concentration. Prior to each tasting, participants rinsed with water, then held each solution in their mouth for about 10 seconds before spitting it out. The recognition threshold was defined as the lowest concentration at which a salty taste was identified, while the preferred concentration corresponded to the solution the participant favored most, following the protocols described by Eriksson et al. (2019) and Tapanee et al. (2021) ^15,16^.

#### 2.5.2 Taste preference and self-reported eating habit

Tomato soup samples were prepared by combining equal parts of tomato passata (Pastificio Riscossa, Corato, Italy; www.riscossa.it) and spring water. Sodium chloride was added to achieve final salt concentrations of 0.15%, 0.30%, 0.50%, 1%, 1.5%, and 2% (w/w). Participants sampled each formulation, rinsing with water between tastings. For each sample, they rated perceived saltiness and palatability using a 100 mm visual analogue scale (VAS), with endpoints labeled “not at all salty” (0%) to “extremely salty” (100%), and “very unpleasant” (0%) to “very pleasant” (100%), respectively.

### 2.6 Clinical and laboratory measurements

Blood pressure (BP) and pulse rate were recorded three times (1-minute intervals) using an Omron HEM-7120 monitor (Omron Healthcare, Japan), with the mean used for analysis. During BP measurement participants sat upright, ensuring their backs rested against the chair and their feet were flat on the ground, with arms aligned to the level of the heart.

#### 2.6.1 Salt Sensitivity of Blood Pressure (SSBP) Assessment

SSBP status for all participants was determined from a prior dietary intervention study within the same cohort, as previously described ^13,14^. In brief, participants were assigned to a high-salt diet (≈9-12 g NaCl/day) followed by a low-salt diet (≈2-3 g NaCl/day) for one week each, with a seven-day washout period. Salt consumption was based on participants’ habitual intake with the prescribed addition or reduction of salt. They were instructed to avoid processed foods to minimize excess salt intake. Participants prepared their own meals without added salt and incorporated the measured salt portions provided, ensuring adherence to the protocol-specified quantities. Twenty-four-hour urine collections were performed at the end of each week to assess adherence, defined as a difference in 24-hour urinary sodium excretion equivalent to more than 100 mmol (approximately 5.8 g of salt) between the two phases. Only participants who were adherent were included in the study.

Ambulatory blood pressure monitoring was performed on the final day of each dietary phase using the CONTEC AMBP70 device (CONTEC Medical Systems Co., Ltd., Qinhuangdao, China). The mean arterial pressure (MAP) was automatically calculated for each phase. SSBP was defined as an increase in MAP of ≥10 mmHg from the low-salt to the high-salt week. Qualified medical personnel from the investigation team monitored each participant’s blood pressure daily during the salt phases through telephone calls. Participants were instructed to measure and report their blood pressure every 30 minutes for 2 hours after each meal. In addition, an alarm was activated for critical blood pressure 0f 170/15 mmHg. No adverse events were reported.

##### 2.6.1.1. Salt Sensitivity Reproducibility

We did not repeat the dietary challenge for the current study, because prior research shows that individual BP responses to salt intake are generally stable over time. Thus, classification from the earlier intervention was used to assign salt-sensitive or salt-resistant status for the current analysis.

Multiple studies indicate that individual blood pressure (BP) responses to salt intake tend to be stable over time. For example, Weinberger (1996) reviewed human salt-sensitivity tests and noted that different methods “have generally been found to be reproducible” ^17^. More recently, Gu et al. (2013) re–tested 487 participants with identical low-sodium/high-sodium diet protocols about 4.5 years apart and found the BP responses to be highly correlated between tests. They conclude that BP responses to dietary sodium are “reproducible and have stable characteristics in the general population” ^18^. In other words, a person classified as salt-sensitive (SS) or salt-resistant (SR) in a prior controlled feeding study is likely to show the same phenotype years later. This evidence supports using a previous dietary salt intervention result to define current salt-sensitivity status without repeating the intervention.

#### 2.6.2 Immediate Pressor Response to Oral Salt (IPROS) Test

An immediate pressor response to oral salt (IPROS) was assessed during this cross-sectional visit. After a baseline BP measurement, participants ingested a solution containing 2 grams of oral salt dissolved in 200 mL of water (NaCl, approximately 34 mmol of sodium) as previously described ^2,19^. Blood pressure was subsequently measured every 10 minutes for 120 minutes. Participants remained seated and rested between measurements. IPROS was defined as a rise in mean arterial pressure (MAP) ≥10 mmHg from baseline at any point during the 120-minute monitoring period. The test was conducted under clinical supervision, and participants were monitored for any adverse effects.

Hypertension was defined according to the JNC 8 (Eighth Joint National Committee) guidelines as a systolic blood pressure ≥140 mmHg and/or diastolic blood pressure ≥90 mmHg, or current use of antihypertensive medication in individuals with a history of elevated blood pressure.

In our study, standard transthoracic echocardiography was performed using M–mode, 2D, and pulsed/tissue Doppler imaging. Linear chamber dimensions, including right ventricular end–diastolic diameter (RVd), left ventricular end–diastolic (LVd) and end–systolic (LVs) diameters, interventricular septal (IVSd) and posterior wall (LVPWd) thicknesses, aortic root (Ao) and left atrial (LA) diameters were measured from parasternal long–axis and apical four–chamber views per American Society of Echocardiography (ASE) guidelines ^20^. Fractional shortening (F/S) was calculated from LVd and LVs, and ejection fraction estimated from LV volumes in the apical two-and four–chamber views. Pulsed–wave Doppler across the mitral valve in the apical four–chamber view provided E and A wave velocities, and the E/A ratio ^20^. While tissue Doppler imaging at the septal annulus yielded e′ to derive the E/e′ ratio. Heart rate was recorded simultaneously, and stroke volume and cardiac output were calculated from LVOT diameter and Doppler velocity time integral using the continuity equation.

Left ventricular hypertrophy was diagnosed by measuring end-diastolic interventricular septal thickness (IVSd) and posterior wall thickness (LVPWd) via parasternal long-axis or 2D–guided M–mode imaging, and calculating LV mass using the Devereux cube formula:

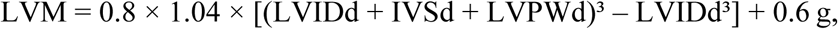

where LVIDd is the LV internal diameter in diastole. LV mass was then indexed (LVMI) to body surface area (BSA=1.73m^2^), with LVH defined using ASE–recommended sex–specific thresholds (LV mass index = LV mass (g) / BSA (m²) > 115 g/m² in men and > 95 g/m² in women)^21,22^.

Body surface area (BSA) was calculated using the Mosteller formula: BSA (m²) = √((Height (cm) and Weight (kg)) / 3600). Relative wall thickness was additionally assessed to characterize concentric (RWT >0.42) versus eccentric (RWT ≤0.42) hypertrophy Relative wall thickness (RWT) was calculated from parasternal long–axis M–mode measurements using the formula: RWT = (2 × posterior wall thickness at end–diastole [PWd]) ÷ left ventricular end–diastolic diameter (LVEDD), following the American Society of Echocardiography guidelines.

Peripheral artery disease (PAD) was diagnosed by measuring the ankle–brachial index (ABI) in the supine position after ≥10 minutes of rest, using a Doppler probe to determine the highest systolic pressures at the dorsalis pedis and posterior tibial arteries and dividing by the higher brachial systolic pressure; an ABI ≤ 0.90 was considered diagnostic of PAD.

Peripheral neuropathy was assessed using biothesiometry: vibration perception threshold (VPT) was measured using the plantar method with a handheld biothesiometer, and neuropathy was defined by VPT ≥ 15 V. Carotid artery evaluation and flow–mediated dilation (FMD) were performed using the cardiovascular suit: carotid intima–media thickness and flow velocities were obtained via duplex ultrasound, and brachial artery FMD was assessed by measuring arterial diameter at baseline and following 5 minutes of forearm cuff occlusion, according to established endothelial function protocols ^23^.

Fasting blood samples were obtained at baseline (prior to salt ingestion) for complete blood count, metabolic, cardiac, renal and inflammatory profiling. We also assessed renal function (serum creatinine) and electrolytes to contextualize salt handling including the renin angiotensin aldosterone system (RAAS). For inflammatory profiling, we measured high-sensitivity C-reactive protein (hsCRP) and key pro-inflammatory cytokines implicated in hypertension: IL-6, tumor necrosis factor-α (TNF-α), and interleukin-17A (IL-17A). Additional cytokines were included as they were relevant in PWH. These were quantified by enzyme-linked immunosorbent assay (ELISA) using commercially available kits, following manufacturer protocols.

The 10–year atherosclerotic cardiovascular disease (ASCVD) risk was calculated using the ACC/AHA Pooled Cohort Equations based on age, sex, race, total and HDL–cholesterol, systolic blood pressure, antihypertensive therapy, smoking status, and diabetes history in individuals aged 40–79 years. The Framingham 2008 general cardiovascular risk score was determined using the sex–specific algorithm incorporating age, total and HDL–cholesterol, systolic blood pressure (treated or untreated), smoking status, and diabetes, to estimate 10–year risk for clinical CVD outcomes.

Estimated glomerular filtration rate (eGFR) was computed from serum creatinine using the CKD–EPI equation (2009), adjusting for age and sex.

### 2.7 Statistical analysis

Data were analyzed using StatCrunch, GraphPad Prism and Python. Continuous variables were summarized as mean ± SD or median (interquartile range) if non-normal. Categorical variables were summarized as counts and percentages. Continuous variables were compared using Welch’s t-tests or Mann-Whitney U tests. Proportions were compared via chi-square tests.

We employed multivariate logistic regression in three primary approaches: 1) analysis of the entire study population adjusted for age, sex, and HIV status; 2) stratified analyses for PWH and PWTH adjusted for age and sex; and 3) a comprehensive model for PWH incorporating additional HIV-specific and cardiovascular variables. Covariates were selected based on their established biological and clinical relevance to SSBP. Multicollinearity was assessed using variance inflation factors (VIF), with no severe multicollinearity detected (VIF < 5 for all variables in final models). This approach was chosen to ensure model parsimony and stability given the sample size. We acknowledge that this strategy does not adjust for all potential confounders and is a limitation of the analysis. p < 0.05 defined significance. Sensitivity analyses for the PWH group, additionally adjusting for HIV-specific factors (ART duration, current CD4+ count, and viral load suppression status), are presented in Supplementary Table S1.

### 2.8 Ethical approval

Prior to commencing the study, ethical approval and administrative clearance were obtained from the National Health Research Authority (NHRA) on 10^th^ of August 2023 (REF:NHREB002/10/08/2023), the Mulungushi University School of Medicine and Health Sciences Research Ethics Committee (MUHSREC) on 08^th^ July 2023 (Ref. No.: SMHS-MU3-2023-004) and renewed on 5^th^ February 2025 (Ref: SMHS-MU2-2023-01), and the Livingstone University Teaching hospital. Written informed consent was obtained from all participants after explaining the study’s objectives, procedures, potential risks, and benefits in a language they understood. Participation was voluntary, and strict measures were taken to uphold data confidentiality and participant privacy throughout the study.

## 3 Results

### 3.1 Participant characteristics

PWH were older (49 ± 12 vs. 44 ± 17 years, p = 0.015) and had lower BMI (25.04 vs. 26.44 kg/m², p= 0.045) and waist circumference (83.79 vs. 87.76 cm, p= 0.037) than PWTH (Fig. 1B–G). Hypertension prevalence did not differ between PWH and PWTH (Fig. 1H), but PWH reported more heart palpitations post-salty meals (Fig. 1K) and higher salt-taste recognition thresholds (0.467 vs. 0.233 g/0.5L, p= 0.0014; Fig. 1L) compared to PWTH. Most PWH used integrase inhibitor-based ART (Fig. 1Q). PWH had higher proportion of individuals with history of tuberculosis (Fig. 1N) and peripheral neuropathy (Fig.1P) compared to PWTH.

### 3.2 Cardiovascular characteristics

Echocardiographic (Fig 2A–K), carotid (Fig 2L–S), and flow-mediated dilation (Fig 2T) parameters did not differ between PWH and PWTH, except for aortic outflow diameter (PWTH: 2.68 ± 2.24 vs. PWH: 2.42 ± 1.63 cm, p< 0.01;Fig 2I) and left atrial diameters (PWTH: 3.22 ± 3.16 vs. PWH: 2.71 ± 0.44 cm, p < 0.01; Fig. 2J), both of which were significantly greater in PWTH than in PWH.

**Figure 2.**
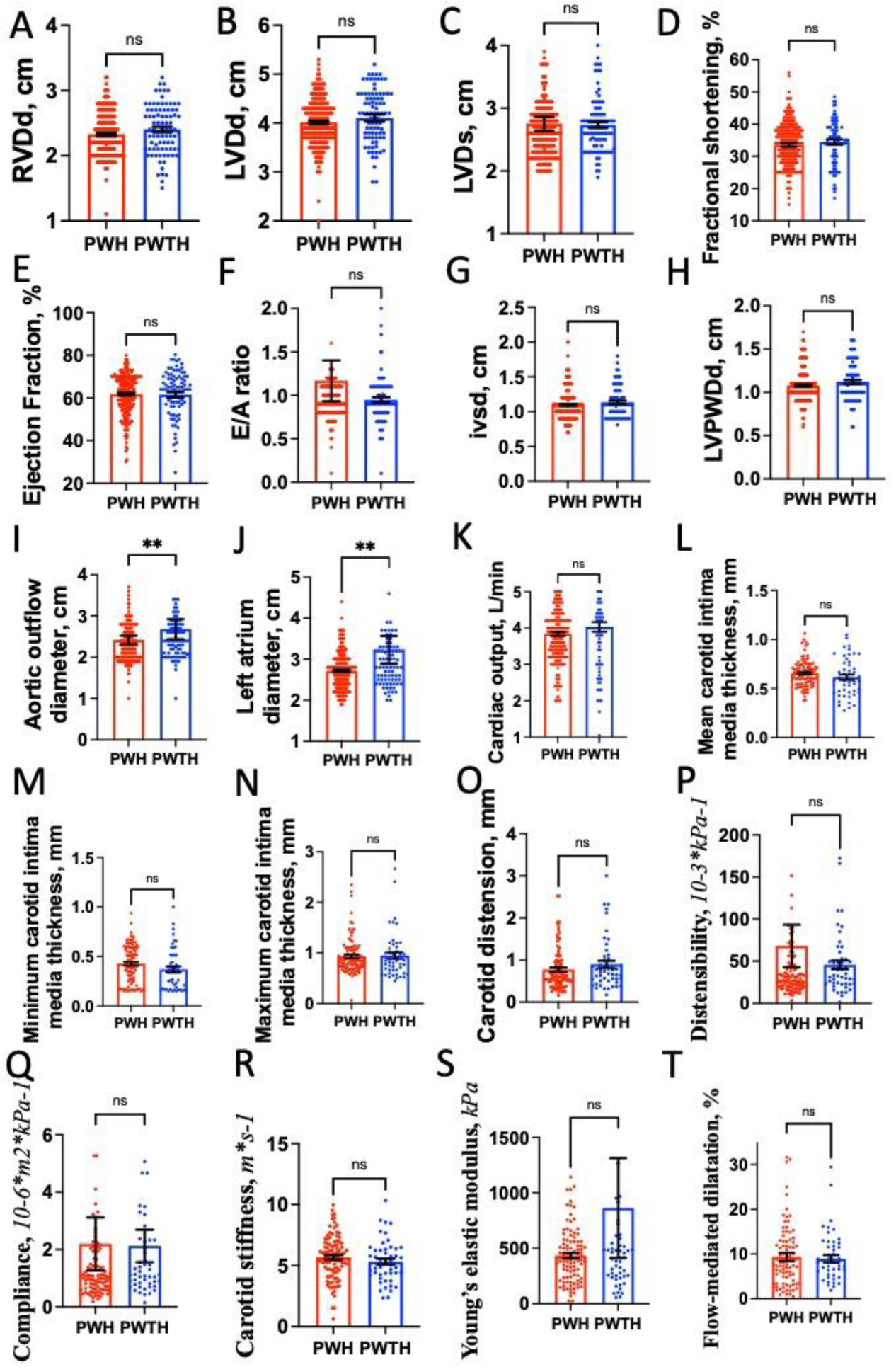
Cardiovascular assessment characteristics between PWH and PWTH. All Echocardiographic (A-K), carotid (L-S) and flow-mediated dilatation (T) parameters were not different between PWH and PWTH except for (I) aortic outflow diameter and (J) left atrial diameters which were higher in PWTH compared to PWH.

### 3.3 Metabolic, Inflammatory, and Renal Profiles

PWH had higher triglyceride (Fig 3B), VLDL-c (Fig 3F), total cholesterol/HDL ratio (Fig 3H), renin (Fig 4C), aldosterone (Fig 4E), angiotensin II (Fig 4D), NT Pro-BNP (Fig 4H), ANP (Fig 4G), d-dimer (Fig 5A), Hs-CRP (Fig 5B), IFN-γ, (Fig 5C), IL-17A (Fig 5D), IgE (Fig 5E), IL-6 (Fig 5F), IL-1 (Fig 5H), IL-5 (Fig 5I), sST2 (Fig 5J), MCH (Fig 6D), WBC (Fig 6E), monocyte count (Fig 6H) eGFR (Fig 6J), uric acid (Fig 6K), plasma chloride (Fig 6N), microalbumin (Fig 6O) and lower HDL (Fig 3C), aldosterone/renin ratio (Fig 4F), RBCs (Fig 6A), plasma sodium (Fig 6L), compared to PWTH, P<0.05.

**Figure 3.**
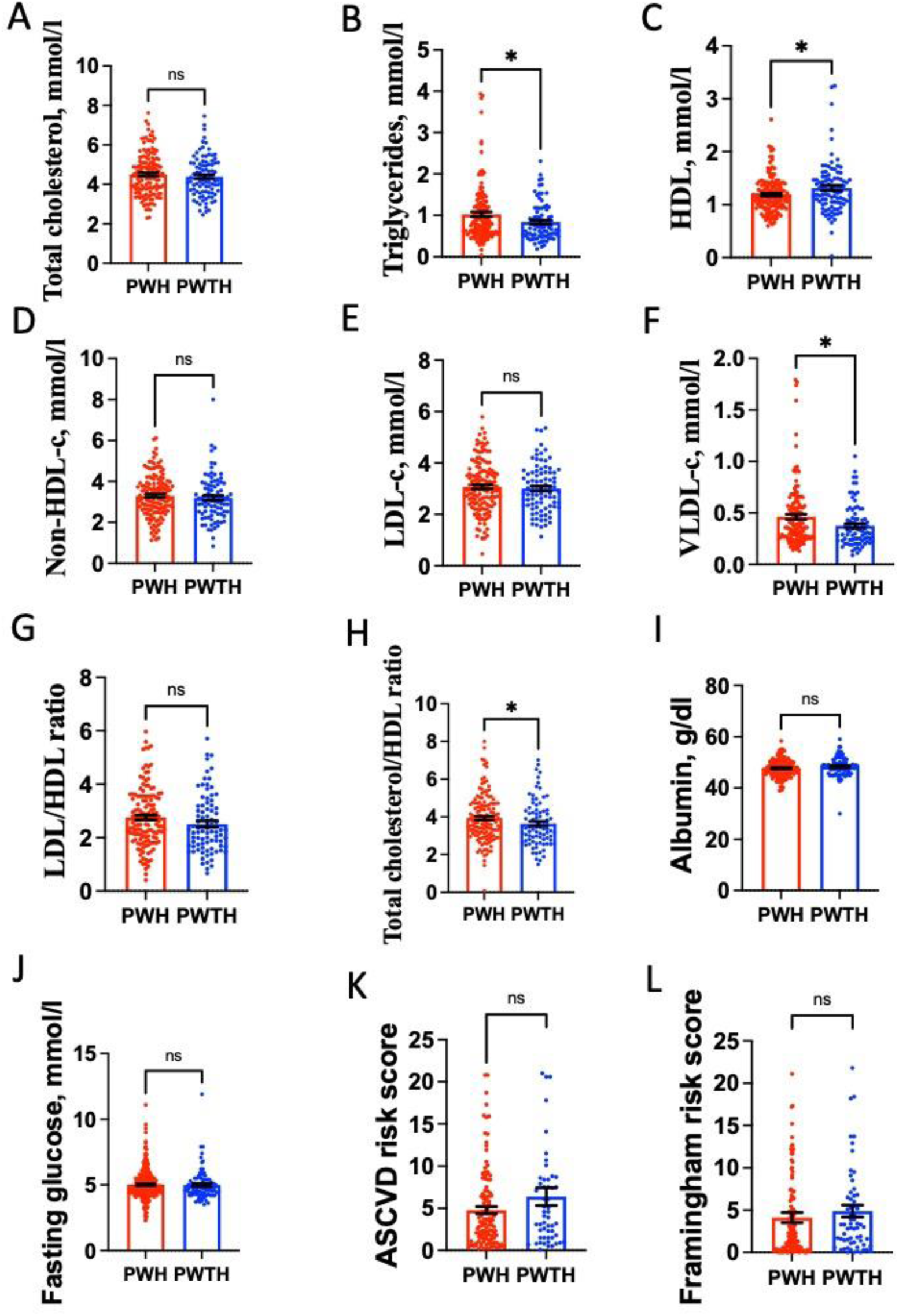
Lipid and metabolic profile between PWH and PWTH. HDL, high density lipoprotein cholesterol; LDL-c, low-density lipoprotein cholesterol; VLDL-c, very low-density lipoprotein cholesterol; ASCVD, atherosclerotic cardiovascular disease risk score

**Figure 4.**
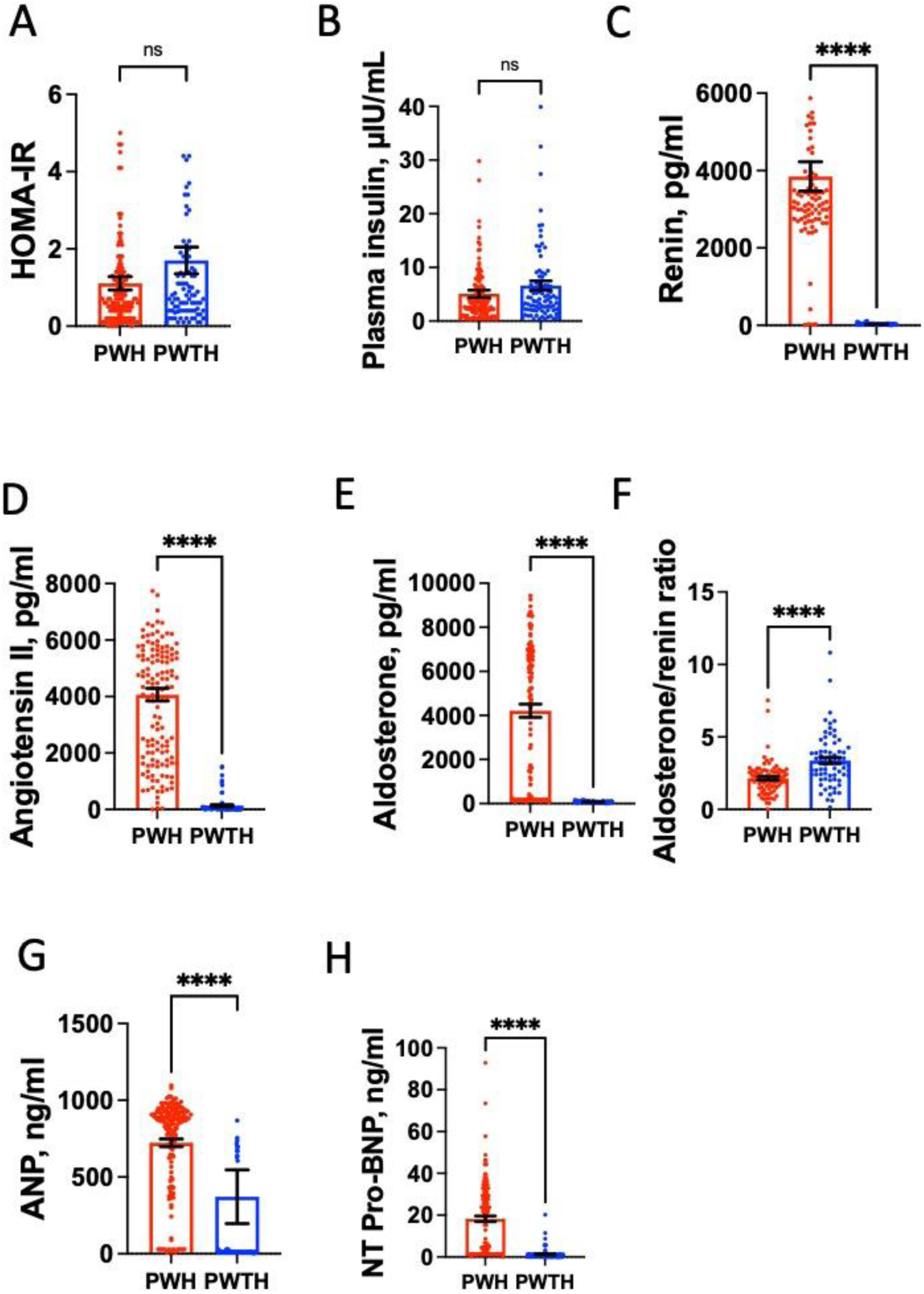
Hormonal profile between PWH and PWTH. ANP, atrial natriuretic peptide; NT-proBNP, N-terminal pro-B-type natriuretic peptide

**Figure 5.**
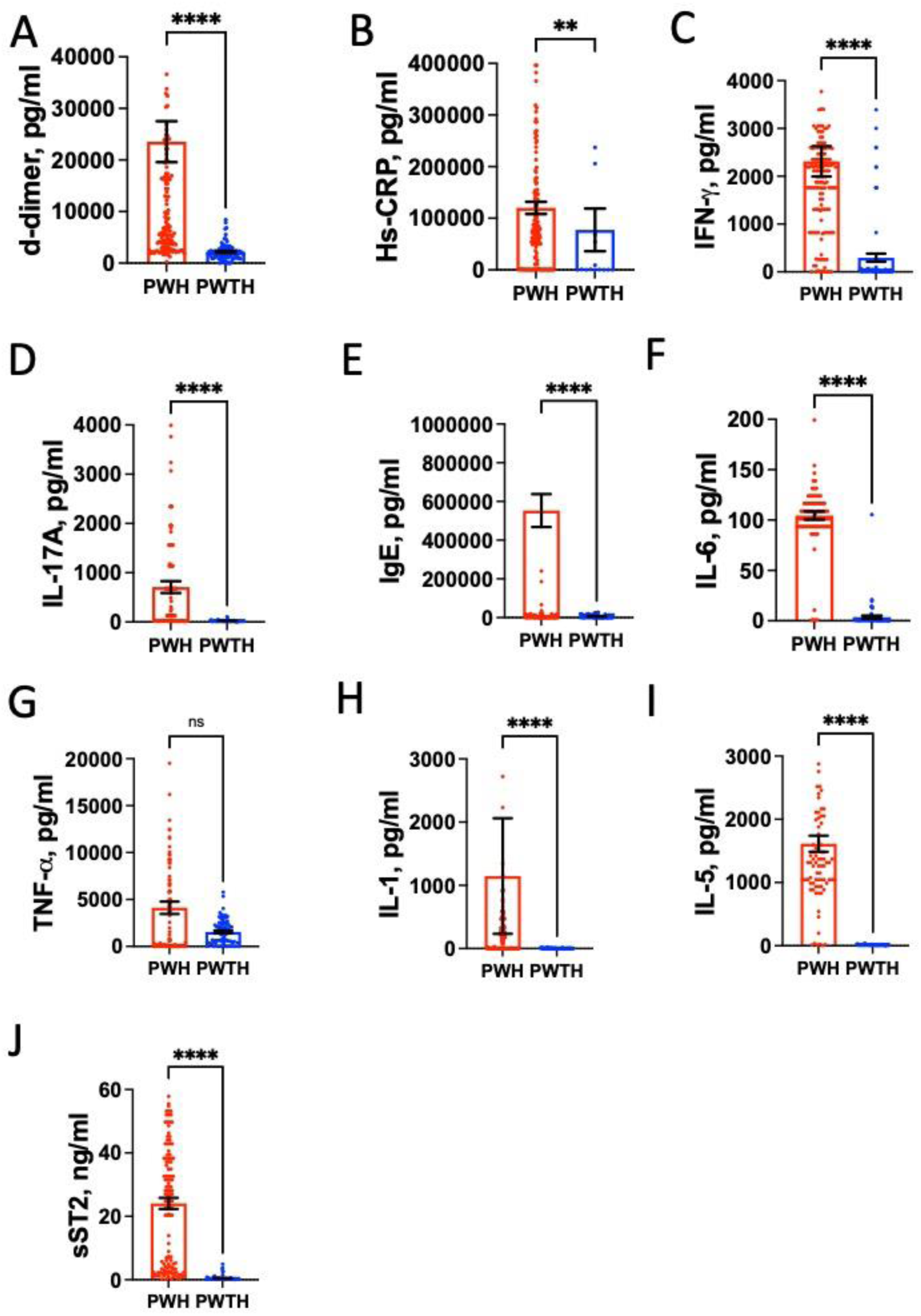
Inflammation markers between PWH and PWTH. D-DIMER, D-dimer (a fibrin degradation product); hsCRP, High-sensitivity C-reactive protein; IFN-γ, Interferon-gamma; IL-17A, Interleukin-17A; IgE, Immunoglobulin E; IL-6, Interleukin-6; TNF-α, Tumor necrosis factor-alpha; IL-1, Interleukin-1; IL-5, Interleukin-5; sST2, Soluble suppression of tumorigenicity 2.

**Figure 6.**
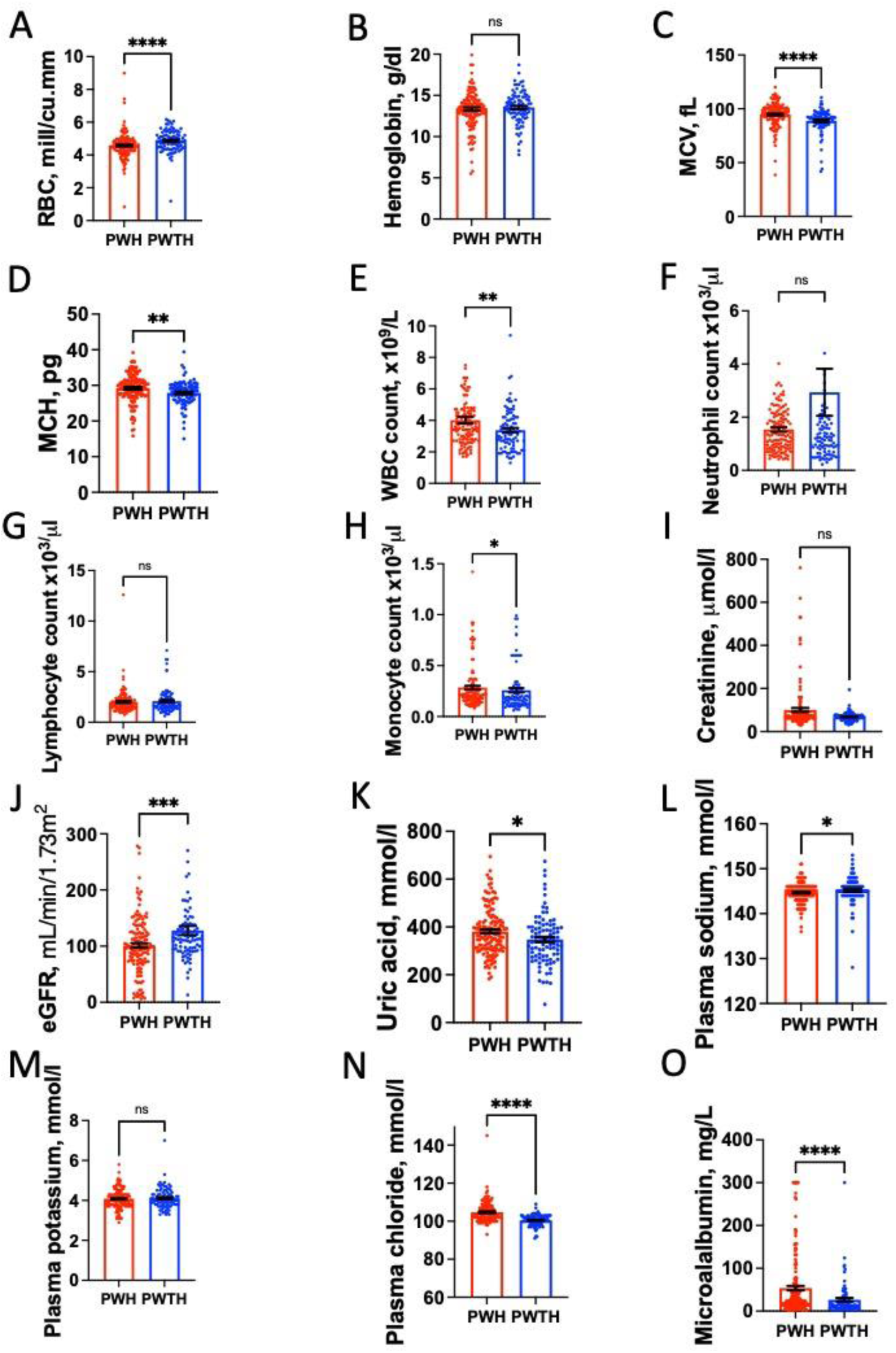
Complete blood count and kidney characteristics between PWH and PWTH. RBC, Red blood cell count; MCV, Mean corpuscular volume; MCH, Mean corpuscular hemoglobin; WBC, White blood cell count; eGFR, Estimated glomerular filtration rate.

#### 3.3.1 Factors associated with SSBP in persons with and without HIV in univariate analysis

Univariate analysis revealed hypertension and an immediate pressor response to oral salt (IPROS) as the strongest correlates of SSBP in both persons with HIV (PWH) and persons without HIV (PWTH) (Table 1). Hypertension demonstrated a markedly elevated odds ratio (OR) in both groups (PWTH: OR = 28.33, 95% CI 7.69–104.35 and PWH: OR = 13.08, 95% CI 6.62–25.81). Similarly, IPROS was a potent correlate for both PWH (OR = 5.41, 95% CI 2.88–10.15) and PWTH (OR = 4.68, 95% CI 1.55–14.07). Among PWH, left ventricular hypertrophy (LVH; OR = 3.90, 95% CI 2.00–7.62) and several cardiac structural markers were strongly associated with SSBP, including relative wall thickness (RWT; OR = 108.50, 95% CI 7.21–1632.63), interventricular septal thickness at end-diastole (IVSd; OR = 38.59, 95% CI 8.30–179.45), left ventricular posterior wall thickness at end-diastole (LVPWd; OR = 58.75, 95% CI 11.20–308.14), and left atrium diameter (OR = 2.87, 95% CI 1.47–5.63). Competency of cardiac valves was also a significant correlate uniquely in PWH (OR = 10.58, 95% CI 2.07–54.05). In contrast, among PWTH, peripheral artery disease (PAD; OR = 6.00, 95% CI 1.91–18.77) and carotid intima-media thickness measures (mean IMT OR = 209.46, 95% CI 2.30–19055.77; min IMT OR = 33.13, 95% CI 1.01–1083.00) showed significant associations. Renal and metabolic factors also exhibited differential associations: plasma creatinine (OR = 1.00 per unit, 95% CI 1.00–1.00) and lower estimated glomerular filtration rate (eGFR; OR = 0.990 per unit, 95% CI 0.981–0.998) were significant correlates only in PWH, while non-HDL cholesterol (OR = 1.69, 95% CI 1.04–2.75) was significant only in PWTH. Atherosclerotic cardiovascular disease (ASCVD) risk score was significantly associated with SSBP in both groups, although the association was stronger in PWH (OR = 1.28, 95% CI 1.14–1.43) compared to PWTH (OR = 1.12, 95% CI 1.01–1.25). Erectile dysfunction (OR = 5.62, 95% CI 1.45–21.80) and a history of tuberculosis (OR = 2.45, 95% CI 1.06–5.65) were additional significant correlates specific to PWH.

**Table 1.**
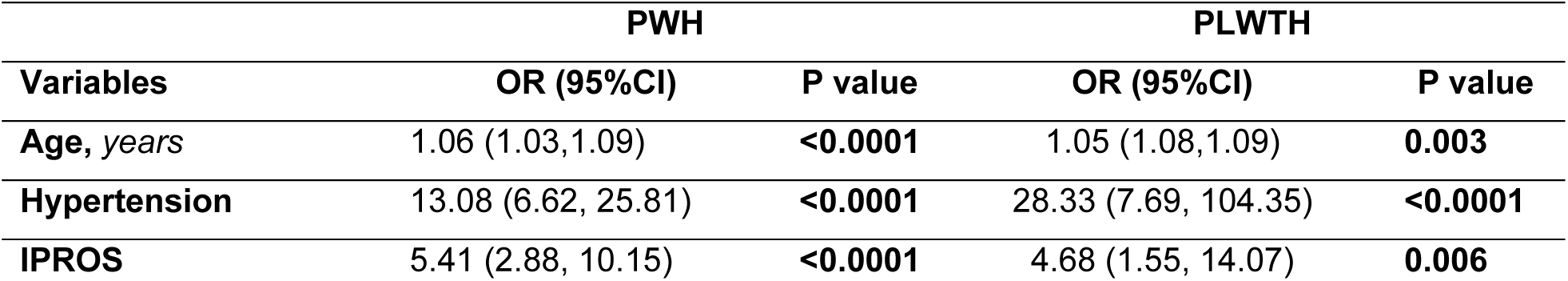
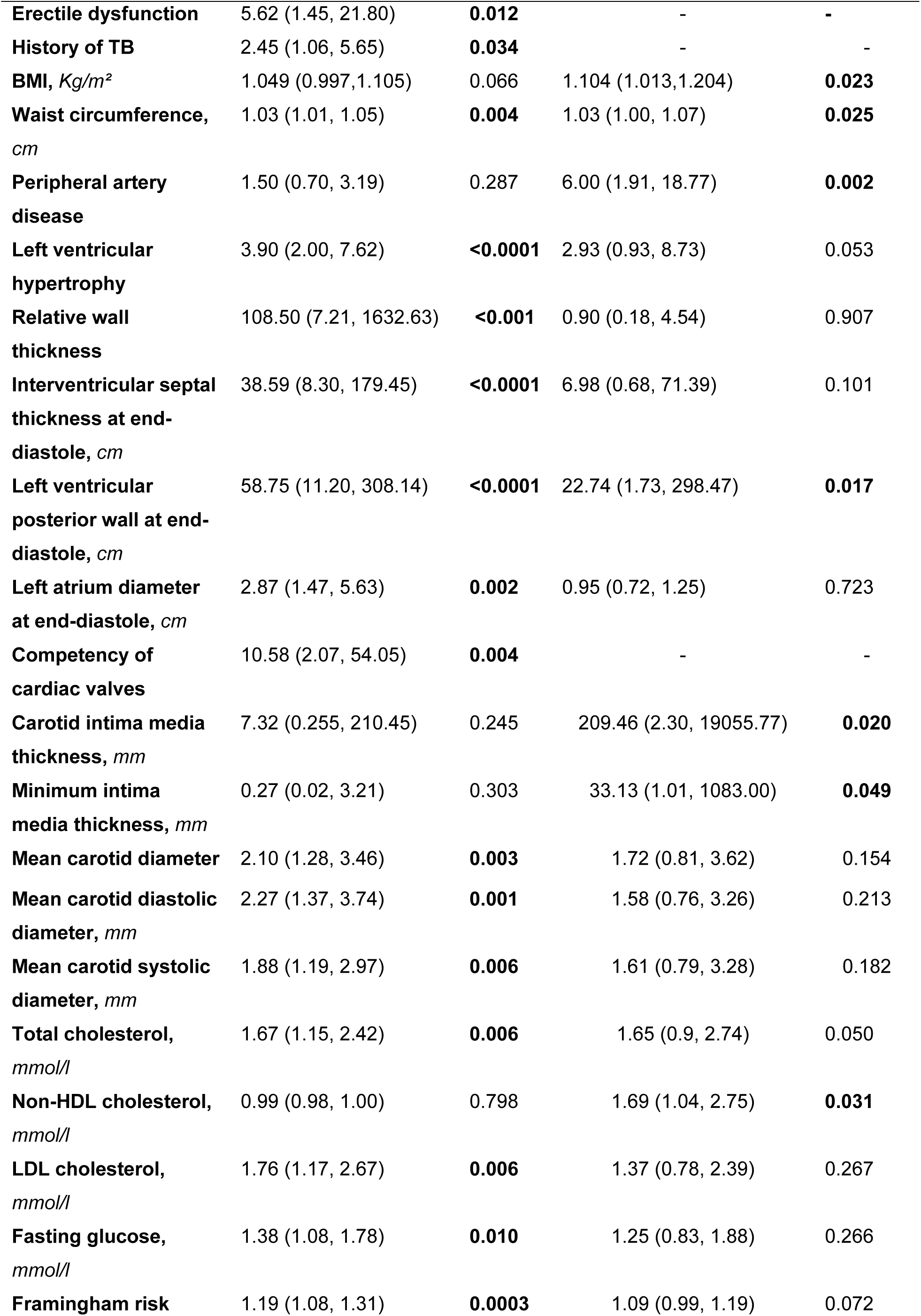
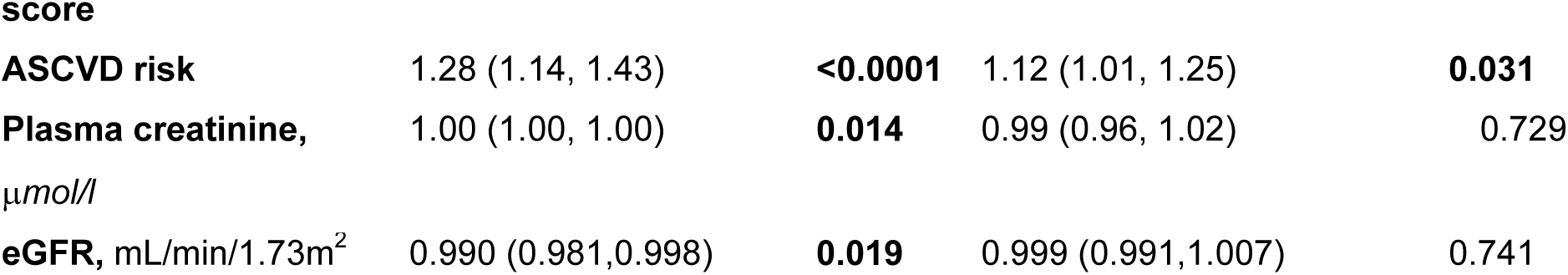
Factors associated with SSBP in persons with and without HIV.

#### 3.3.2 Factors associated with SSBP in the whole study population adjusted for age, HIV and sex in Multivariate analysis

A multivariate analysis of the entire study population, adjusted for age, HIV status, and sex, identified several factors independently associated with SSBP (Table 2). Hypertension was the strongest independent predictor, with hypertensive individuals having over twelve times the odds of having SSBP compared to normotensive individuals (Adjusted Odds Ratio [AOR] = 12.28, 95% CI 6.46–23.35, p < 0.0001). Cardiovascular and metabolic markers were also significant independent correlates. A positive IPROS test was strongly associated with SSBP (AOR = 5.70, 95% CI 3.18–10.21, p < 0.0001). Similarly, the presence of Peripheral Artery Disease (AOR = 2.63, 95% CI 1.36–5.10, p = 0.004) and higher calculated cardiovascular risk scores, including the Framingham Risk Score (AOR = 1.15 per unit, 95% CI 1.04–1.28, p = 0.0064) and the Atherosclerotic Cardiovascular Disease (ASCVD) Risk Score (AOR = 1.38 per unit, 95% CI 1.16–1.64, p = 0.0002), were all independently associated with increased odds of SSBP.

**Table 2.**
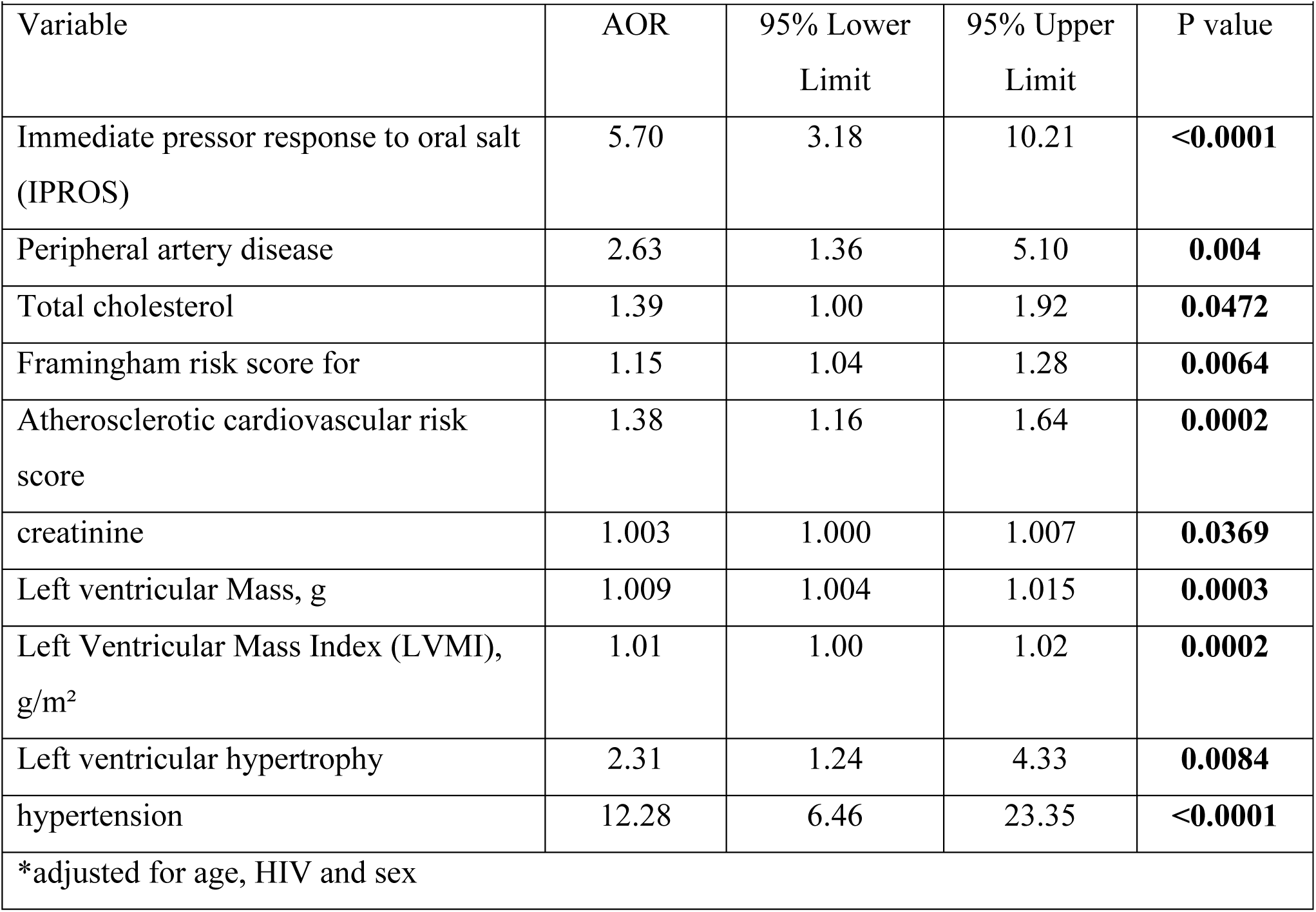
Significant Factors associated with SSBP in the whole study population in Multivariate analysis model.

|  |  |  |  |  |
| --- | --- | --- | --- | --- |
| Variable | AOR | 95% Lower Limit | 95% Upper Limit | P value |
| Immediate pressor response to oral salt (IPROS) | 5.70 | 3.18 | 10.21 | **<0.0001** |
| Peripheral artery disease | 2.63 | 1.36 | 5.10 | **0.004** |
| Total cholesterol | 1.39 | 1.00 | 1.92 | **0.0472** |
| Framingham risk score for | 1.15 | 1.04 | 1.28 | **0.0064** |
| Atherosclerotic cardiovascular risk score | 1.38 | 1.16 | 1.64 | **0.0002** |
| creatinine | 1.003 | 1.000 | 1.007 | **0.0369** |
| Left ventricular Mass, g | 1.009 | 1.004 | 1.015 | **0.0003** |
| Left Ventricular Mass Index (LVMI), g/m<sup>2</sup> | 1.01 | 1.00 | 1.02 | **0.0002** |
| Left ventricular hypertrophy | 2.31 | 1.24 | 4.33 | **0.0084** |
| hypertension | 12.28 | 6.46 | 23.35 | **<0.0001** |
| *adjusted for age, HIV and sex |  |  |  |  |

Cardiac structure and renal function further demonstrated independent associations. Indicators of left ventricular remodeling and hypertrophy were significant predictors, including Left Ventricular Mass (AOR = 1.009 per unit, 95% CI 1.004–1.015, p = 0.0003), Left Ventricular Mass Index (LVMI; AOR = 1.01 per unit, 95% CI 1.00–1.02, p = 0.0002), and the presence of Left Ventricular Hypertrophy (LVH) itself (AOR = 2.31, 95% CI 1.24–4.33, p = 0.0084). Additionally, higher plasma creatinine levels were independently associated with SSBP (AOR = 1.003 per µmol/L, 95% CI 1.000–1.007, p = 0.0369). Finally, a higher total cholesterol level also showed a significant, albeit weaker, independent association with SSBP (AOR = 1.39 per unit, 95% CI 1.00–1.92, p = 0.0472).

In summary, after adjusting for age, HIV status, and sex, SSBP in this cohort was independently associated with a cluster of factors encompassing hypertension, acute salt sensitivity (IPROS), peripheral vascular disease, elevated cardiovascular risk scores, adverse cardiac remodeling, and subtle renal impairment.

#### 3.3.3 Factors associated with SSBP in persons with and without HIV in multivariate analysis

Multivariate analysis model 1, adjusting for age and sex, identified distinct independent predictors of SSBP for PWH and PWTH (Table 3). Hypertension was the strongest correlate, with adjusted odds ratios (AOR) of 10.27 (95% CI 5.02–21.00) for PWH and 24.68 (95% CI 5.45–111.73) for PWTH. IPROS remained a robust independent correlate in both groups, with adjusted odds ratios (AOR) of 6.24 (95% CI 3.16–12.32) for PWH and 4.73 (95% CI 1.47–15.21) for PWTH.

**Table 3.** Factors associated with SSBP in multivariate analysis model 1*.

| Variables | PLWH |  | PLWTH |  |
| --- | --- | --- | --- | --- |
|  | AOR (95%CI) | P value | AOR (95%CI) | P value |
| Immediate Pressor Response to Oral Salt (IPROS) | 6.24 (3.16, 12.32) | **<0.0001** | 4.73 (1.47, 15.21) | **0.009** |
| Married | 2.61 (1.28, 5.30) | **0.0078** | 0.48 (0.13, 1.67) | 0.252 |
| Hypertensive crisis on ambulatory blood pressure | 18.15 (.39, 47.78) | **0.0199** | - |  |
| Controlled blood pressure | 1.42 (0.34, 5.82) | 0.620 | 0.02 (0.00, 0.56) | **0.024** |
| Peripheral artery disease | 1.87 (0.84, 4.17) | 0.124 | 7.15 (1.96, 26.05) | **0.0028** |
| Interventricular Septal Thickness in Diastole (IVSDd), cm | 19.06 (3.95, 91.84) | **0.0002** | 1.46 (0.09, 22.14) | 0.7826 |
| Left Ventricular Posterior Wall in Diastole (LVPWd), cm | 24.25 (4.25, 138.32) | **0.0003** | 4.71 (0.17, 126.13) | 0.3549 |
| Left Ventricular Mass Index (LVMI), g/m<sup>2</sup> | 1.02 (1.01, 1.03) | **0.0002** | 1.01 (0.99, 1.02) | 0.299 |
| Left ventricular mass (LVM), g | 1.01 (1.00, 1.02) | **0.0002** | 1.00 (0.99, 1.01) | 0.320 |
| Left ventricular hypertrophy | 2.75 (1.33, 5.68) | **0.0006** | 1.40 (0.39, 4.94) | 0.0600 |
| Relative Wall Thickness (RWT) | 30.52 (1.90, 488.18) | **0.0156** | 0.44 (0.00, 39.86) | 0.721 |
| Carotid mean diameter, mm | 2.07 (1.24, 3.46) | **0.0052** | 1.14 (0.64, 2.02) | 0.6517 |
| Carotid diameter in diastole, mm | 2.26 (1.33, 3.83) | **0.0024** | 0.83 (0.34, 2.03) | 0.6886 |
| Carotid diameter in systole, mm | 1.88 (1.17, 3.00) | **0.0079** | 1.05 (0.46, 2.40) | 0.9019 |
| Framingham risk score | 1.23 (1.05, 1.44) | **0.0077** | 1.08 (0.94, 1.23) | 0.232 |
| Atherosclerotic Cardiovascular Disease (ASCVD) Risk Score | 1.59 (1.24, 2.03) | **0.0002** | 1.15 (0.93, 1.42) | 0.189 |
| Plasma creatinine | 1.003 (1.000, 1.007) | **0.037** | 0.99 (0.96, 1.03) | 0.654 |
| Hypertension | 10.27 (5.02, 21.00) | **<0.0001** | 24.68 (5.45, 111.73) | **<0.0001** |

\*Adjusted for age and sex. AOR: Adjusted Odds Ratio; CI: Confidence Interval.

**Table 4.**
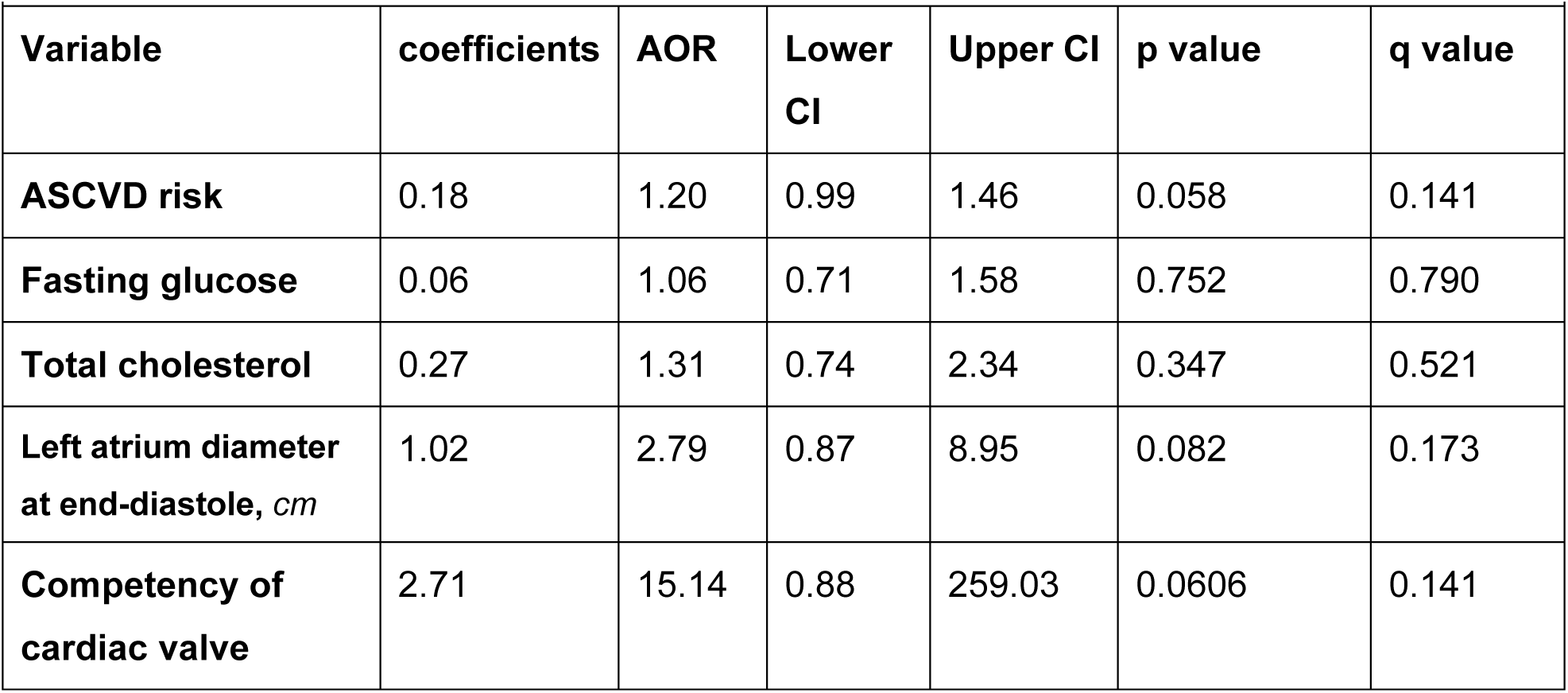
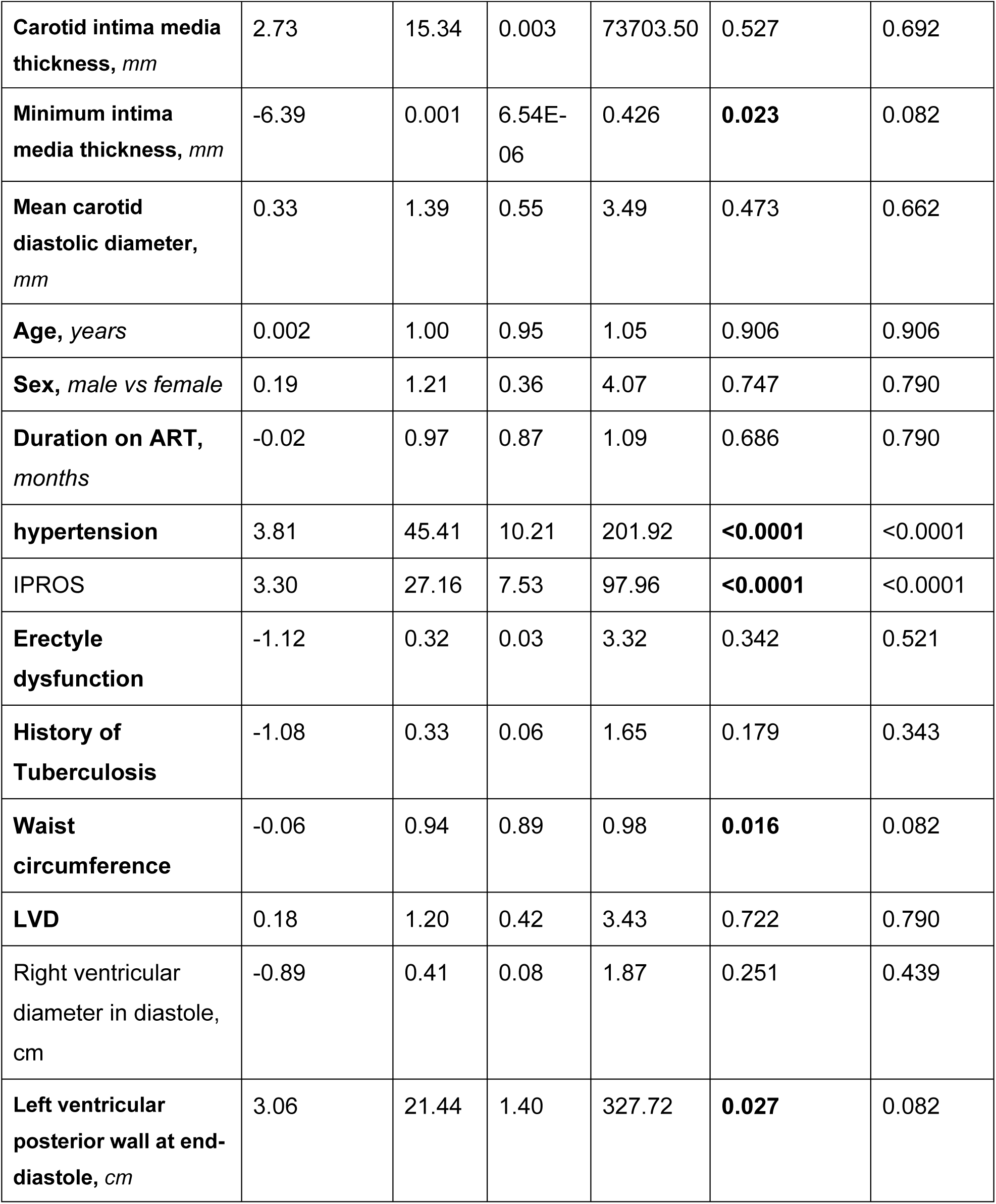
Factors associated with SSBP in multivariate analysis model 2.

Among PWH, several cardiac structural markers were independently associated with SSBP. These included greater interventricular septal thickness at end-diastole (IVSd; AOR = 19.06, 95% CI 3.95–91.84), left ventricular posterior wall thickness at end-diastole (LVPWd; AOR = 24.25, 95% CI 4.25–138.32), left ventricular mass index (LVMI; AOR = 1.02 per unit, 95% CI 1.01–1.03), and the presence of left ventricular hypertrophy (LVH; AOR = 2.75, 95% CI 1.33–5.68). Relative wall thickness (RWT; AOR = 30.52, 95% CI 1.90–488.18), carotid diameters, Framingham risk score (AOR = 1.23, 95% CI 1.05–1.44), ASCVD risk score (AOR = 1.59, 95% CI 1.24–2.03), and plasma creatinine (AOR = 1.003 per µmol/L, 95% CI 1.000–1.007) were also independent correlates specific to PWH. Additionally, being married was associated with higher odds of SSBP in PWH (AOR = 2.61, 95% CI 1.28–5.30).

Among PWTH, the independent correlates of SSBP were more limited. Peripheral artery disease was a strong predictor (AOR = 7.15, 95% CI 1.96–26.05), while having controlled blood pressure was associated with markedly reduced odds of SSBP (AOR = 0.02, 95% CI 0.00–0.56). Notably, the cardiac structural markers and risk scores that were significant in PWH were not independently associated with SSBP in PWTH.

#### 3.3.3 Multivariate Analysis of Expanded Covariates for SSBP in PWH

For purposes of incorporating all covariates, we tested for multicollinearity for the PWH and results were acceptable based on variance inflation factor as no severe inflation was detected among the included correlates (Supplementary Table S1). The Best multivariable model 2 (Table 3) built on the L1-selected set revealed CV AUC ≈ 0.892 suggesting strong discrimination for SSBP among HIV-positive participants. The summary table containing coefficient estimates, standard errors, z-stats, and p-values for interpretability are found in Supplementary Table S2.

A more comprehensive multivariate logistic regression model 2 constructed to identify independent factors associated with SSBP in PWH, incorporating a wider array of demographic, clinical, cardiovascular, and HIV-specific variables (Figure 7 and Table 3). After adjusting for all other covariates in the model, hypertension and an IPROS remained the most robust and statistically significant correlates of SSBP.

**Figure 7.**
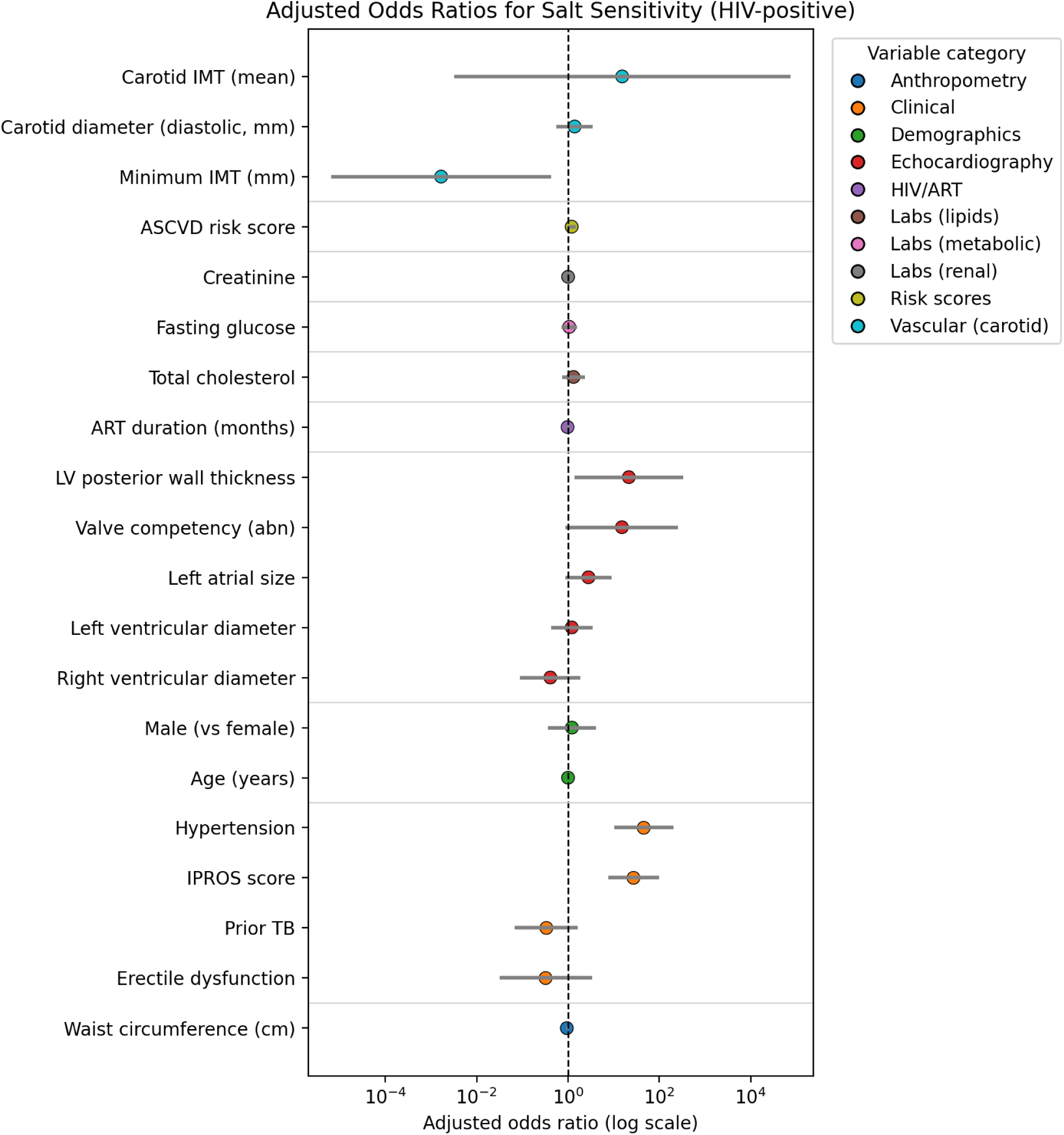
Forest plot of odds ratios for multiple variables and their association with Salt sensitivity of blood pressure. TB, tuberculosis; IMT, intima media thickness; ASCVD, atherosclerotic cardiovascular disease; LV, left ventricle; abn, abnormal; IPROS, immediate pressor response to oral salt.

The presence of hypertension was associated with a dramatically increased odds of SSBP (AOR = 45.41, 95% CI 10.21–201.92, p < 0.0001), Table 3. Similarly, a positive IPROS test was strongly associated with SSBP (AOR = 27.16, 95% CI 7.53–97.96, p < 0.0001). These associations held strong after correction for multiple testing (q < 0.0001).

Several other factors demonstrated notable associations, though they did not retain formal statistical significance after multiple testing correction (q-value > 0.05). Left ventricular posterior wall thickness at end-diastole (LVPWd) was associated with higher odds of SSBP (AOR = 21.44, 95% CI 1.40–327.72, p = 0.027). Conversely, a larger waist circumference was associated with a reduced odds of SSBP (AOR = 0.94 per cm, 95% CI 0.89–0.98, p = 0.016). A thinner minimum carotid intima-media thickness was also associated with lower odds of SSBP (AOR = 0.001, 95% CI 6.54E-06–0.426, p = 0.023). Trends were observed for ASCVD risk score (AOR = 1.20, p = 0.058), left atrium diameter (AOR = 2.79, p = 0.082), and cardiac valve competency (AOR = 15.14, p = 0.061), though these were not statistically significant.

Notably, duration on antiretroviral therapy (ART) (AOR = 0.97 per month, p = 0.686) was not a significant independent predictor of SSBP in this model. Other factors, including age, sex, fasting glucose, total cholesterol, and history of tuberculosis, were also not independently associated with SSBP after multivariate adjustment.

### 3.4 Discussion

This comprehensive study provides novel insights into the determinants of salt-sensitive blood pressure (SSBP) in Zambian adults with HIV (PWH), revealing distinct pathophysiological mechanisms that differentiate them from their HIV-negative counterparts (PWTH). Our findings underscore the profound impact of chronic inflammation and metabolic dysregulation on sodium handling and cardiovascular risk in this population, while also highlighting the clinical utility of the immediate pressor response to oral salt (IPROS) as a pragmatic biomarker. Below, we contextualize these findings within the broader scientific literature, discuss their clinical implications, and outline the strengths and limitations of our work.

## Key Findings and Integration with Existing Literature

PWH exhibited significantly elevated inflammatory markers, renin-angiotensin-aldosterone system (RAAS) activation (higher renin, aldosterone, angiotensin II), and altered natriuretic peptide responses (NT-proBNP, ANP) compared to PWTH. These findings align with established models of HIV-associated cardiovascular disease, where persistent immune activation driven by viral reservoirs, microbial translocation, and antiretroviral therapy (ART) promotes endothelial dysfunction and oxidative stress ^8,24^. Crucially, hypertension and IPROS emerged as dominant predictors of SSBP in both groups, but their impact was magnified in PWH (IPROS AOR = 19.90 vs. 18.49 in PWTH). This suggests that HIV-specific factors, such as monocyte activation and inflammasome priming, amplify salt-induced endothelial injury ^8,24^. The paradoxical observation of RAAS activation coexisting with lower plasma sodium in PWH further indicates HIV-related renal tubular dysfunction, potentially due to inflammation-driven sodium retention ^25,26^.

Divergent SSBP correlates between groups were particularly striking. PWH demonstrated strong associations between SSBP and cardiac remodeling (LV mass index), renal impairment (plasma creatinine), and calculated ASCVD risk. These findings extend our prior work showing nocturnal non-dipping hypertension in Zambian PWH ^19^, suggesting that subclinical organ damage exacerbates salt sensitivity in this population. PWTH relied more on traditional pathways, with atrial natriuretic peptide (ANP) and peripheral artery disease as primary SSBP correlates. This aligns with population-based studies where ANP serves as a compensatory mechanism for sodium handling ^27–29^.

Notably, the heightened salt-taste recognition thresholds in PWH may reflect HIV-related sensory neuropathy or inflammation-mediated alterations in gustatory signaling. This novel finding complements our recent work on erythrocyte glycocalyx sensitivity in salt-sensitive women as well as the role of HIV in taste sensitivity, suggesting a bidirectional relationship between sodium intake and vascular dysfunction ^2,30^. However, the underlying mechanisms for altered taste perception remain unclear and future studies should include genetic analyses of taste receptor genes and more detailed assessments of sensory neuropathy to elucidate these mechanisms.

Our multivariate analyses reveal distinct patterns of SSBP correlates between PWH and PWTH, with PWH demonstrating a more complex profile involving cardiac structural changes, renal parameters, and some novel factors. The association between marital status and SSBP in PWH is particularly intriguing and warrants further investigation into potential psychosocial, behavioral, or socioeconomic mediators of this relationship. The comprehensive analysis of the entire cohort identified hypertension, IPROS, peripheral artery disease, left ventricular hypertrophy, and cardiovascular risk scores as independent correlates of SSBP, underscoring the multifactorial nature of salt sensitivity that transcends HIV status. However, the stratified analyses revealed that the pathophysiology of SSBP in PWH involves more extensive end-organ damage, as evidenced by the multiple cardiac structural parameters (IVSd, LVPWd, LVMI, RWT) that remained significant after multivariate adjustment.

Our expanded multivariate model (Model 2) revealed novel, independent associations beyond the dominant correlates of hypertension and IPROS. The finding that a larger waist circumference was protective against SSBP was unexpected and warrants further investigation into the potential role of body composition and adipokine signaling in sodium homeostasis in PWH. Conversely, the strong association between increased left ventricular posterior wall thickness (LVPWd) and SSBP underscores the intimate link between salt-sensitive pathways and the development of concentric cardiac remodeling, a known precursor to heart failure with preserved ejection fraction.

## Clinical Implications: Bridging Mechanisms to Management

Our multivariate analysis revealed that IPROS, a dynamic marker of acute endothelial stress, strongly predicted SSBP. The consistent independent association of IPROS with SSBP across all analytical approaches (whole population, stratified, and comprehensive PWH models) strongly supports its validation as a practical clinical tool for identifying salt-sensitive individuals in resource-limited settings, where complex SSBP protocols are impractical. A systolic BP rise ≥10 mmHg at 30 minutes post-salt load could identify high-risk individuals warranting aggressive dietary sodium restriction (<2 g/day) or RAAS inhibitor therapy ^13,19^. Furthermore, the prominent role of cardiac structural parameters in PWH suggests that echocardiographic assessment may be particularly valuable for risk stratification in this population.

In addition, ASCVD risk scores outperformed traditional hypertension metrics in predicting SSBP in PWH, underscoring the need for integrated risk assessment tools that incorporate HIV-specific factors (e.g., CD4 nadir, ART duration). Early echocardiography to detect LV hypertrophy may also refine risk stratification.

### Strengths of the Study

This investigation offers several significant advancements:

Comprehensive Phenotyping: As the first African HIV cohort with concurrent SSBP, dietary, sensory, and cardiometabolic-inflammatory assessments, we identified novel links (e.g., taste perception, IPROS) previously unexplored in this population. Methodological Rigor: Our use of standardized SSBP protocols, echocardiography, and ASCVD scoring enhances cross-study comparability. The inclusion of both normotensive and hypertensive participants allowed dissection of HIV-specific drivers independent of BP status. Contextual Relevance: By focusing on a Zambian cohort, where high-salt diets and HIV prevalence intersect, our findings address a critical gap in global cardiovascular health equity. The predominance of integrase inhibitor-based ART (Fig. 1Q) also reflects contemporary treatment patterns, enhancing real-world applicability. Translational Biomarkers: Our study also offers hope on the use of IPROS as a feasible clinic-based tool, bridging mechanistic research and clinical practice in low-resource settings ^19^.

### Limitations

Several constraints merit consideration. Firstly, the cross-sectional design precludes causal inferences between HIV, inflammation, and SSBP. Longitudinal studies tracking IPROS, inflammatory markers, and end-organ damage progression are needed to establish causality. Secondly, ART Homogeneity: Most participants used integrase inhibitors, limiting exploration of how protease inhibitor-based regimens, which affect lipid metabolism, influence SSBP. Thirdly, salt-sensitivity status was taken from a prior dietary protocol and not reassessed. We based classification on the earlier intervention because studies indicate salt-sensitivity is a relatively stable phenotype over time ^17,18^. However, salt-sensitivity can vary, and without re-testing some individuals may be misclassified. This should be noted when interpreting the findings. Fourthly, mechanistic gaps: While inflammatory markers were elevated, direct assays of endothelial glycocalyx integrity or sodium buffering capacity were not performed ^2^. Future research should incorporate these direct measures to provide a more comprehensive understanding of the vascular mechanisms involved. Another important limitation is generalizability. Our findings may not extend to PWH with uncontrolled viremia, diabetes, or advanced renal disease, as these were exclusion criteria. The inclusion of participants with these comorbidities in future studies would improve the generalizability of the findings and allow assessment of their impact on salt sensitivity. In addition to the previously noted limitations, we acknowledge that the unexpected association between marital status and SSBP in PWH, while statistically significant, requires cautious interpretation and validation in future studies. The psychological and behavioral mechanisms underlying this association remain speculative at this stage.

### 3.5 Conclusion

In this Zambian cohort, multivariate analyses revealed that SSBP correlates differ substantially between PWH and PWTH, with PWH exhibiting a more complex profile involving cardiac structural changes, and renal parameters. PWH exhibited distinct metabolic-inflammatory signatures associated with SSBP, with IPROS and hypertension as pivotal predictors. The consistent strong association of IPROS with SSBP across all analytical models supports its potential as a practical clinical screening tool. Clinically, IPROS assessment offers a pragmatic tool for identifying high-risk individuals, while our data on lipid mediators and taste perception open new avenues for targeted interventions. HIV is associated with amplified salt sensitivity via chronic inflammation, RAAS dysregulation, and renal-cardiac remodeling, extending findings from our prior work. The distinct correlates in PWH highlight the need for HIV-specific approaches to salt sensitivity assessment and management. Furthermore, the identification of waist circumference and left ventricular posterior wall thickness as independent correlates highlights novel pathways involving body composition and cardiac structure that may contribute to SSBP in this population. Future studies should explore anti-inflammatory strategies and should also include more in-depth mechanistic studies investigating renal sodium handling and immune activation to better understand how these pathways drive salt-sensitive hypertension in PWH. Given the rising burden of hypertension-related morbidity in sub-Saharan Africa, these findings underscore the urgency of integrating HIV status into cardiovascular risk paradigms.

## 5 Author Contributions

Writing original draft- SKM and JPP. Revising drafts - CAG, LP, AK. Conceptualization and data curation- SKM, AK and LP. Formal analysis, visualization, validation – AK, JPP, CAG, LP, SKM. Funding acquisition – SKM and AK

## Data Availability

The data that support the findings of this study are not publicly available due to ethical restrictions and the need to protect participant confidentiality. De-identified data are available from the corresponding author upon reasonable request, subject to a data use agreement and approval from the Mulungushi University School of Medicine and Health Sciences Research Ethics Committee.

## Acknowledgments

We thank participants, clinic staff, HAND Research group research assistants and the Livingstone Centre for Prevention and Translational Science.

